# Improving Offloading in Diabetic Feet: 3D-Printed Custom Accommodative Insoles Outperform Standard of Care

**DOI:** 10.1101/2025.08.07.25333211

**Authors:** Brittney C. Muir, Kimberly A. Nickerson, Christina Carranza, Mathew Sunil Varre, Avocet Nagle-Christensen, Dylan Heino, Ellen Li, William R. Ledoux, Scott Telfer

**Affiliations:** VA RR&D Center for Limb Loss and MoBility (CLiMB), VA Puget Sound Health Care System, 1660 S Columbian Way, MS 151, Seattle, WA 98108; Department of Mechanical Engineering, University of Washington, 3900 E Stevens Way NE, Box 352600, Seattle, WA 98195; Department of Orthopaedic Surgery and Sports Medicine, University of Washington, 1959 NE Pacific St., Box 356500, Seattle, WA, United States, 98195

**Author notes:** Corresponding Author: Brittney Muir, Ph.D. Address: Center for Limb Loss and MoBility (CLiMB) VA Puget Sound Health Care System 1660 South Columbian Way MS 151 Seattle, WA 98108 USA. **Twitter Summary:** 3D-printed custom accommodative insoles reduce plantar pressure more effectively than standard care, offering a promising personalized solution for diabetic foot offloading and complication prevention.

**Keywords:** diabetic insoles, 3D printed diabetic insoles, plantar pressure, pressure time integral

## Abstract

**Objective:** 3D-printed custom accommodative insoles may offer improved plantar pressure offloading compared to standard-of-care insoles for individuals with diabetes. We have developed two versions of 3D-printed accommodative insoles that have patient-specific geometries as well as patient-specific stiffnesses and structural behaviors. This study evaluated two 3D-printed custom accommodative insoles against standard-of-care and no insole.

**Research Design and Methods:** This in-laboratory clinical trial enrolled 25 adults with diabetes and high plantar pressure. Participants completed over-ground walking trials under the following conditions: no insole, standard-of-care insoles, pressure-based insoles, and finite element insoles. Peak plantar pressure and pressure-time integral were measured in high-risk regions of interest and adjacent regions. Participant comfort and preference were recorded.

**Results:** All insoles significantly reduced peak pressure and pressure-time integral in the regions of interest compared to no insole (p<0.001) by greater than 14%. Both pressure-based and finite element insoles further reduced peak pressure (22% and 24% respectively) and pressure-time integral (19% and 35% respectively) beyond standard-of-care (p<0.001). Finite element insoles reduced pressure-time integral and peak pressure in adjacent regions (p<0.01), indicating broader offloading. While finite element insoles demonstrated strong biomechanical performance, participant preferences and feedback indicated improvements in fit and comfort are needed across all insoles before clinical implementation.

Conclusions

This study demonstrates the potential of personalized, 3D-printed custom accommodative insoles as a patient-centered approach to offloading plantar pressure in individuals with diabetes. These technologies may support more effective and tailored strategies in preventing diabetic foot complications. Further research will assess long-term outcomes and optimize comfort and fit.

**Highlights:**

- Why did we undertake this study? Individuals with diabetes and high plantar pressure are at risk for foot complications, and current insoles may not provide sufficient pressure relief.
- What is the specific question(s) we wanted to answer? Can two types of 3D-printed custom accommodative insoles—pressure-based and finite element designs—provide superior plantar pressure offloading compared to standard-of-care and no insole conditions?
- What did we find? Both 3D-printed insole types significantly outperformed standard-of-care insoles in reducing peak plantar pressure and pressure-time integral in the region of interest. Finite element designs also reduced pressure in adjacent regions.
- What are the implications of our findings? Personalized 3D-printed insoles may improve diabetic foot care by offering targeted offloading. Optimizing fit and comfort is needed before broader clinical use.

## INTRODUCTION

Diabetic foot ulcers are a major cause of lower limb amputations and impose a significant economic burden on healthcare systems (1,2). Treating a diabetic foot ulcer can cost up to $85,000, (2) and the five-year mortality rate after ulceration is 30% (3). Currently, more than 38.4 million Americans, or 11.6% of the population, live with diabetes, (4). In 2016, more than 130,000 diabetic-related amputations were performed in the U.S. (5). Preventing foot ulceration in people with diabetes is essential to reduce amputation rates (6,7).

Custom accommodative insoles are prescribed to patients with diabetes who are at risk of developing plantar ulcers (8,9). Custom insoles aim to reduce plantar pressures with a weight-bearing surface that conforms to the patient’s foot. The current standard of care (SoC) insoles are designed to match a patient’s foot shape, captured via a foam crush box, and are typically constructed from a milled EVA foam block then covered with a uniform sheet of PORON® and plastazote foam (10). The material properties of these insoles are generic, which may limit the clinician’s ability to optimize the device’s pressure-reducing capability at the patient-specific level.

Advances in 3D printing techniques and the associated materials and software have enabled researchers to create new latticed 3D printed metamaterials where mechanical properties are derived not only from the base material but also from the lattice microstructures within the metamaterial. Our group has recently developed a novel digital workflow to design and 3D print custom accommodative insoles that incorporate foot shape and plantar pressure (11–13). These insoles not only have patient-specific geometry but also patient-specific stiffness and structural behavior. Our previous work has indicated that our 3D-printed custom accommodative insoles with in-shoe pressure-based offloading regions (PB-CAI) are effective in alleviating areas of high plantar pressure in healthy participants (11) showing a statistically significant 14% reduction in peak plantar pressure in key areas compared to the standard of care. While SoC and various custom accommodative insole designs, including PB-CAI insoles, have been shown to be effective in reducing the plantar pressure in the forefoot region (8,11,14–16), a more rigorous evaluation of our 3D printed insoles is needed in the target population, individuals with diabetes and high plantar pressures.

Beyond these advances, the inclusion of computational simulation methods such as finite element (FE) analysis into the design process for offloading insoles has been proposed as an optimization technique to produce high-performing offloading insoles without the need for multiple physical iterations of the device (FE-CAI). To this end, we developed an approach that uses simplified models and accessible measurement technologies to overcome some of the complexity and time-consuming nature of running these types of simulations (17) and demonstrated its effectiveness for the design of offloading insoles (18).

Our prior work has described the insole design workflow and mechanical testing of the 3D printed material in depth (11–13,17,18). The purpose of this study is to determine if our two styles of novel 3D printed insoles can be designed to provide greater reductions in plantar pressures for at-risk areas of the foot than the SoC insoles. Plantar pressure reduction with the SoC and a no insole (NI) condition were compared to two types of 3D printed diabetic insoles 1) a PB-CAI and 2) a FE-CA (Figure 1, Top). We hypothesize that 1) the SoC will reduce the peak plantar pressure (PPP) and pressure-time integral (PTI) compared to NI. 2) Both 3D printed insoles (PB-CAI and FE-CAI) will reduce PPP and PTI compared to the SoC. 3) The FE-CAI will have the greatest reduction in PPP and PTI compared to the other insoles.

**Figure 1.**
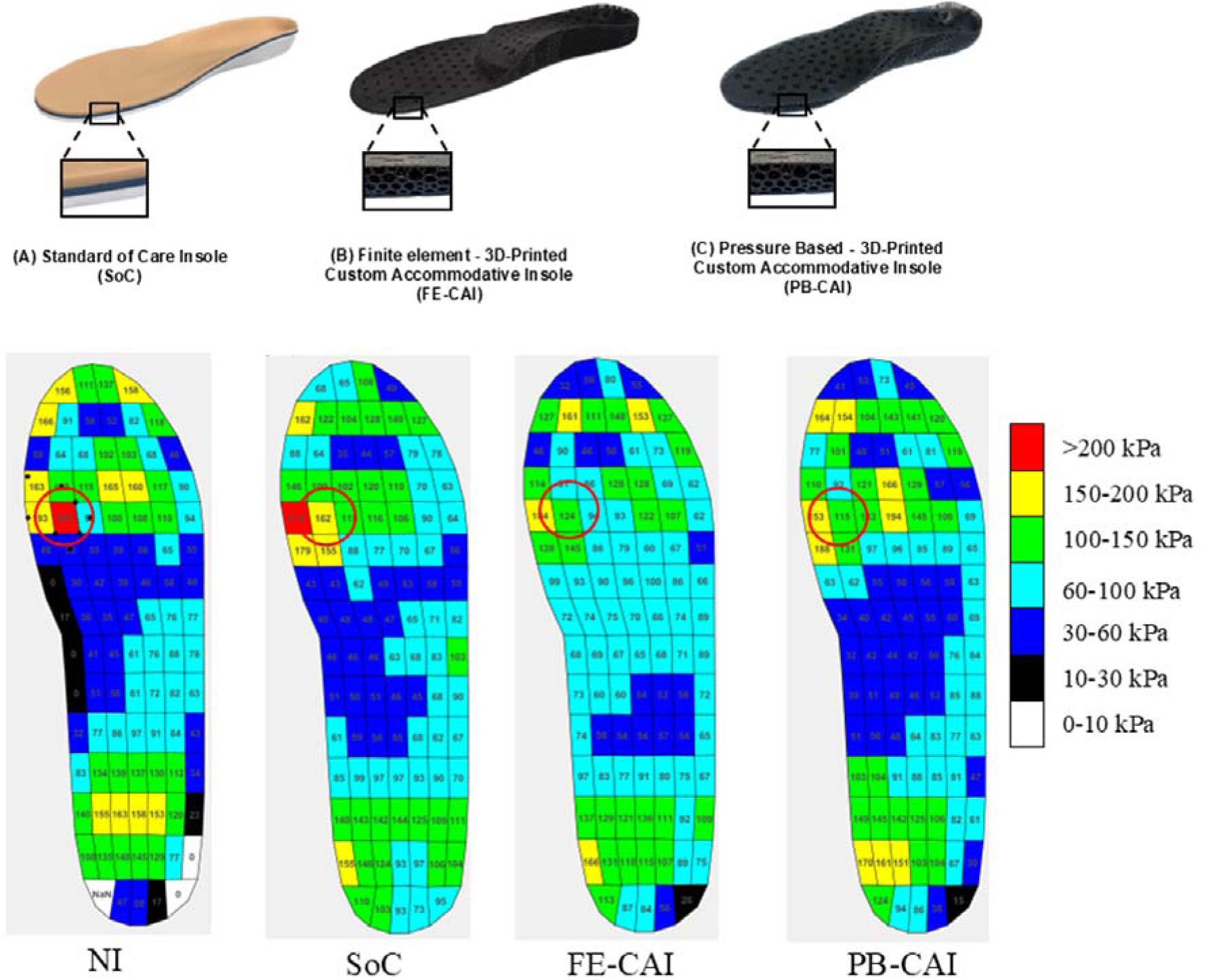
(Top) The three types of insoles that were fabricated. A) Standard of Care insole (SoC). B) Finite Element 3D-Printed Custom Accommodative Insole (FE-CAI). C) Pressure-Based 3D Printed Custom Accommodative Insole (PB-CAI). (Bottom) A representative peak plantar pressure distribution of a participant during shod walking in the four insole conditions.

## METHODS

### Participants

Twenty-five participants diagnosed with diabetes (24 males, Age (years):73.6 ± 6.3, height (cm): 174.8 ± 6.2) were recruited for the study. The inclusion criteria were adults diagnosed with diabetes, high plantar pressure, no history of ulceration, and who had the ability to walk independently across an 8-meter-long walkway without an assistive device. Written informed consent of the participants was obtained before they participated in this Department of Veterans Affairs Puget Sound Institutional Review Board-approved study (IRB# 1588070).

### First visit

Participant demographic and health information were collected. A foot assessment was performed by a certified prosthetist–orthotist and foam crushbox impressions of the participant’s feet were obtained. Measurements of forefoot anatomy and tissue deformation were captured using an Aixplorer ultrasound machine with an SL 18-5 transducer (SuperSonic Imagine, Aix en Provence, France) as described in (18). Participants were then fitted with standardized research shoes (Dr. Comfort, WI, USA) and in-shoe pedar-x (novel GmbH, Munich, DE) pressure-measuring insoles and completed five successful self-selected speed walking trials along an 8-meter walkway. This was considered the NI condition. Finally, participants completed 10 (five with each foot) barefoot walking pressure scans using the emed pressure system (novel GmbH, Munich, DE). Walking speed was recorded using infrared timing gates and monitored to ensure the difference in walking speeds between trials was less than 10%.

### Insole design

Three insole types were generated: 1) SoC, 2) PB-CAI, and 3) FE-CAI.

The SoC insole (Figure 1 Top A) was fabricated using typical methods described in our previous work (13). The foam crush box was shipped to a local contracted clinical insole manufacturer. The insole manufacturer used the information prescribed by the orthotist to fabricate the SoC insoles that consisted of an EVA base with a 4-mm Plastazote-PORON foam top layer (Hudak et al., 2022); this is the most commonly prescribed material combination for diabetic insoles.

Two pairs of 3D printed insoles, FE-CAI (Figure 1 Top B) and PB-CAI (Figure 1 Top C), were generated following our design and manufacturing workflow described in Hudak et al. (13). Briefly, the insoles were designed using the same foam crush box impression used in the SoC insole. The foam crush boxes were 3D scanned (GoScan3d, Creaform, Levis, Quebec, Canada) and the areas with high plantar pressure in the forefoot were identified using the in-shoe and barefoot pressure data and a custom MATLAB script (The MathWorks Inc., Natick, Massachusetts). Computer models of the insole were generated using Rhino v7 (Robert McNeel & Associates, WA, USA) and the foam crush box impressions and areas of high pressure of the participants were combined. The models were sent to Carbon 3D (Carbon Inc., Redwood City, CA, USA) to be latticed and printed using their Elastomeric Polyurethane (EPU) 41 material. Fabrication details for each insole are described below.

The design of the 3D printed insole varies based on the method employed to offload the areas of high pressure. The design process for the PB-CAI has been described in detail elsewhere (11–13). Briefly, regions of high pressure were defined as in-shoe peak pressure of 200 kPa or more (19–21) and a sparse “soft” lattice metamaterial was printed in locations identified as experiencing high pressure. A dual-layered lattice metamaterial, comprised of a rigid bottom layer and a softer, accommodative top layer, was designed with Carbon’s internal software to match the compressive stiffnesses replicating the SoC (12) was printed in the rest of the insole volume. A 4 mm Voronoi cell was used for each lattice layer, with strut diameters of 1.03 mm and 0.68 mm for the rigid and accommodative layers, respectively. The offloading region had a single strut diameter of 0.6 mm.

The design process for the FE-CAI has been described in detail elsewhere (17,18). Briefly, a basic insole design is produced based on shape and pressure data. The design includes the notable feature of a metatarsal bar that is defined by the proximal forefoot pressure region contour. A geometrically simplified version of the forefoot based on the foam box geometry, ultrasound measurements of tissue thickness, materials properties, and metatarsal geometry along with the corresponding region of the insole are imported into the FE software FEBio Studio (22). The material properties of the insole are determined from mechanical testing (12). A simulation is run with metatarsal forces matching the peak forefoot loads, defined from the barefoot pressure data, and if the predicted pressure for any metatarsal head region is above 200kPa, the metatarsal bar height in the insole design is increased and the simulation repeated until a “safe” load is predicted. A 4 mm Voronoi cell lattice was used, but with a uniform-layer design was created with strut diameter of 0.85 mm (equal to the average of the accommodating and rigid layer struts in the PB-CAI).

### Second Visit

In-shoe plantar pressures were recorded using the standardized research shoe and pedar-x insoles while the participants completed the five walking trials in each of the insole conditions: SoC, PB-CAI, and FE-CAI, lasting up to 15 minutes per condition. The order of the insole intervention tested was randomized and blinded from the participant to avoid bias until the data collection session concluded. Finally, qualitative questions regarding ease of use and comfort of the devices were asked, and responses were recorded. These questions include “What did you like about this insole?”, “What did you dislike about this insole?”, and “Which one of the insoles did you like the best/least and why?”

### Data Analysis

The plantar pressure data from the second visit for the four insole conditions was used for further analysis. Plantar pressure data for each walking trial was processed using an open-source software package (*pressuRe*) which is built for the R statistical computing environment and designed to process, analyze, and visualize pressure data (23). The peak pressure of each sensor cell (sensel) was identified for each step, and then the peak pressures were averaged across all steps. PTI was calculated following the method described in (24). The region of interest (ROI) was defined as the region(s) with in-shoe peak pressure of ≥200 kPa at the initial visit. Maximum peak pressure was defined as the highest peak pressure in the ROI for each participant (e.g., red circle in Figure 1 Bottom) and the adjacent (ADJ; within one sensel of the ROI) region. The adjacent region was selected for analysis as we did not want to reduce pressure in the ROI to increase pressure in the area adjacent. Left and right steps were included in the data analysis. A one-way ANOVA, conducted in R, was used to determine if there were differences in PPP or PTI (the dependent variable) by insole type (NI, SoC, PB-CAI, and FE-CAI) within each location in the insole (ROI vs. ADJ). Alpha was set to 0.05. Tukey post hoc pairwise comparisons were conducted to determine group differences (Table 1). Means, standard deviations (Figure 2), and percentage change (Table 1) for each dependent variable by insole combination were calculated. Success rates for reducing on keeping PPP to below 200 kPa in the ROI and ADJ were calculated for each insole condition. User feedback for likes and dislikes of the insoles were transcribed and grouped into themes that emerged from the qualitative responses (25).

**Figure 2.**
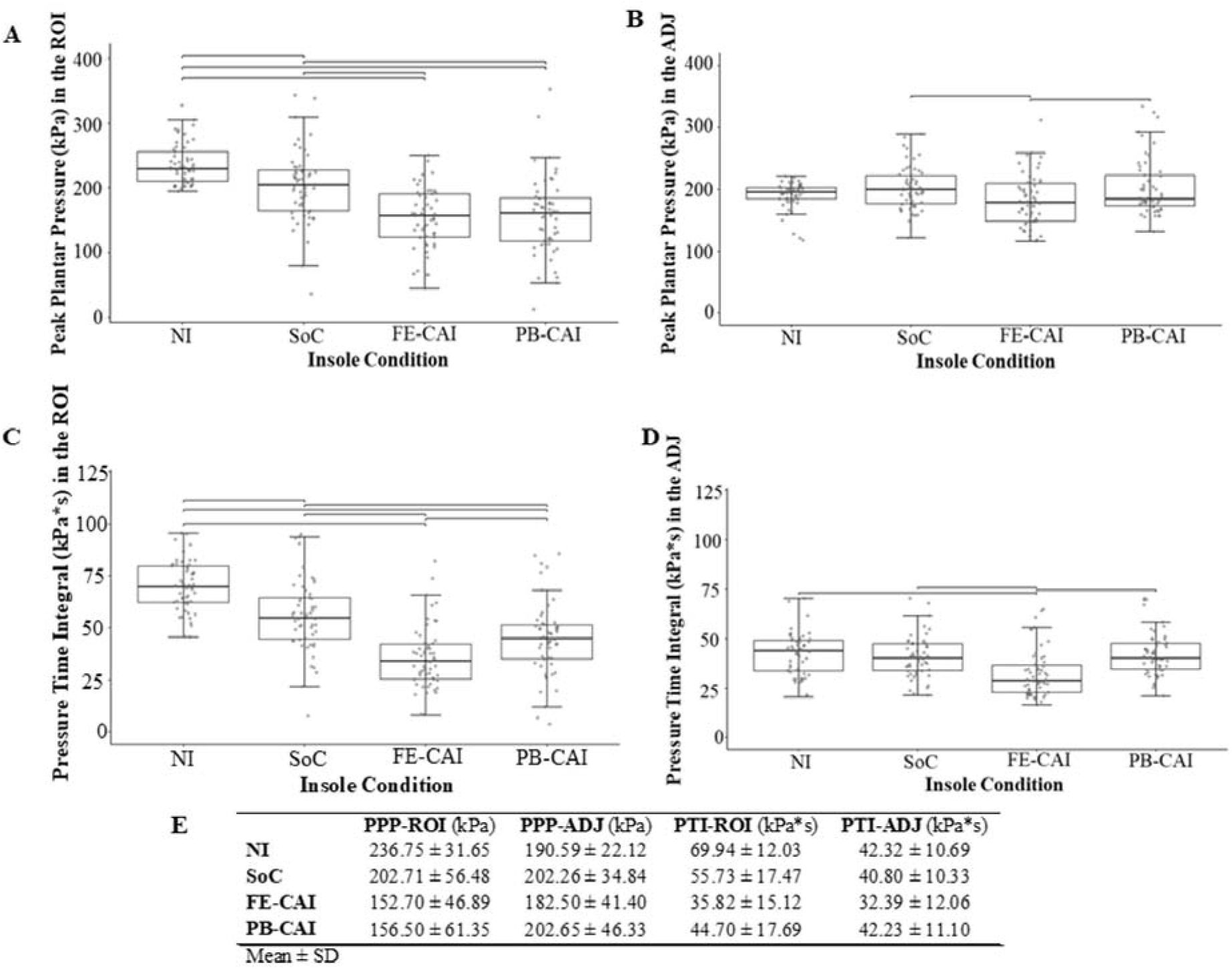
Peak plantar pressure during walking in the region of interest (ROI) (a) and the adjacent (ADJ) (b), and pressure time integral in the ROI (c) and the ADJ (d) across the four insole conditions. Significance bars indicate significant post hoc comparisons, p-value < 0.05. (E) Means and standard deviations expressed as a function of insole by dependent variable.

**Table 1.** Post hoc analysis p-values and 95% confidence intervals and percentage change in PPP and PTI. Negative values indicate a reduction in pressure from the first condition to the second condition.

|  | PPP-ROI |  |  | PPP-ADJ |  |  | PTI-ROI |  |  | PTI-ADJ |  |  |
| --- | --- | --- | --- | --- | --- | --- | --- | --- | --- | --- | --- | --- |
|  | p-value | 95% CI | % change | p-value | 95% CI | % change | p-value | 95% CI | % change | p-value | 95% CI | % change |
| NI - SoC | **0.0035** | -59.42, -8.65 | -14 | 0.3650 | -6.89, 30.23 | 6 | **<0.001** | -22.09, -6.31 | 6 | 0.894 | -7.06, 4.02 | -3 |
| NI- FE-CAI | **<0.001** | -109.44, -58.67 | -35 | 0.6721 | -26.65, 10.47 | -4 | **<0.001** | -42.00, -26.22 | -4 | **<0.001** | -15.46, -4.38 | -23 |
| NI - PB-CAI | **<0.001** | -105.63, -54.87 | -33 | 0.3401 | -6.59, 30.70 | 6 | **<0.001** | -33.13, -17.34 | 6 | 1.000 | -5.63, 5.45 | 0 |
| SoC - FE-CAI | **<0.001** | -75.40, -24.63 | -24 | **0.0318** | -38.32, -1.21 | -9 | **<0.001** | -27.77, -12.06 | -9 | **0.0024** | -13.92, -2.89 | -20 |
| SoC - PB-CAI | **<0.001** | -71.60, -20.83 | -22 | 0.9999 | -18.26, 19.03 | 0 | **0.0019** | -18.89, -3.18 | 0 | 0.908 | -4.08, 6.94 | 3 |
| FE-CAI - PB-CAI | 0.9800 | -21.58, 29.19 | 2 | **0.0285** | 1.50, 38.79 | 11 | **0.0198** | 1.02, 16.73 | 11 | **<0.001** | 4.33, 15.35 | 30 |

## RESULTS

### Plantar Pressure

*PPP in ROI* demonstrated a significant effect of insole (F(3, 212) = 28.90, p<0.001, η²=0.29; Figure 2A). Post hoc analysis indicated that PPP was reduced in the ROI for all insole versions compared to the NI condition (p<0.01), and FE-CAI and PB-CAI showed a further significant decrease in PPP compared to the SoC (p<0.001). FE-CAI and PB-CAI were not significantly different from each other (p=0.9). The NI, SOC, FE-CAI, and PB-CAI had a PPP in the ROI below 200 kPa in 6%, 48%, 87%, and 81% of instances, respectively.

*PPP in ADJ* demonstrated a significant effect of insole (F(3, 211) = 3.69, p=0.013, η²=0.0.5; Figure 2B). Post hoc analysis indicated that PPP in ADJ was significantly lower in the FE-CAI (<0.031) compared to the NI and PB-CAI. There were no other significant differences (p>0.341). Please note that the ROI was defined using a different software package from the initial in-shoe pressure data than the main analysis, leading to some variation in the steps selected at each stage and some ADJ regions showing values slightly over 200kPa (Figure 2). The NI, SOC, FE-CAI, and PB-CAI had a PPP in the ADJ below 200 kPa in 59%, 54%, 69%, and 65% of instances, respectively.

*PTI in ROI* demonstrated a significant effect of insole (F(3, 211) = 46.64, p<0.001, η²=0.4; Figure 2C). Post hoc analysis indicated that PTI in ROI was significantly reduced with all versions of the insoles compared to the NI condition (p<0.001), and FE-CAI and PB-CAI showed a further significant decrease in PTI compared to the SoC (p<0.002), and FE-CAI showed a further reduction in PTI compared to PB-CAI (p=0.019).

*PTI in ADJ* demonstrated a significant effect of insole (F(3, 211) = 9.91, p<0.001, η²=0.1; Figure 2D). Post hoc analysis indicated that PTI in ADJ was significantly reduced with the FE-CAI compared to all other conditions (p<0.01). No other significant differences were observed (p>0.9).

### Insole preference

When asked to select their favorite insole, 40% (10/25) of participants selected the PB-CAI as their favorite, 36% (9/25) of participants selected the SoC as their favorite, and 24% (6/25) of participants selected the FE-CAI as their favorite. These preferences were primarily driven by comfort/pain, with the primary factors being supportiveness (specifically arch support), stability/balance, materials, and fit in the shoe (Table 2).

**Table 2.** Insole preferences.

|  |  |  |  |
| --- | --- | --- | --- |
| Likes |  |  |  |
|  | **SoC** | **PB-CAI** | **FE-CAI** |
|  | *Comfort/ Pain* |  |  |
|  | • Comfortable<br>• Feels like no insole at all<br>• Balls for feet feel engaged with surface | • Comfortable<br>• No discomfort<br>• Could wear all day<br>• Feel like I walk better in these<br>• Not painful | • No pain |
|  | *Supportiveness* |  |  |
|  | • Supportive | • Support in arch area<br>• Whole foot felt supported<br>• Supportive | • Very supportive<br>• Support through arch<br>• |
|  | *Stability/ Balance* |  |  |
|  | • Stable in arch<br>• Balance is more centered | • Balance seems better<br>• Improved stability | • More stable |
|  | *Materials* |  |  |
|  | • Really bouncy, lots of cushion<br>• Like memory style foam insole |  |  |
|  | *Fit in the shoe* |  |  |
|  | • Snug fitting | • Didn’t need much adjustment |  |
|  | *Misc.* |  |  |
|  |  |  | • Might like more with time<br>• Helped my feet<br>• Better than own insoles<br>• More formed to foot |
| Dislikes |  |  |  |
|  | **SoC** | **PB-CAI** | **FE-CAI** |
|  | • Felt a little lumpy | • More irritation<br>• Lateral R heel painful<br>• Does no think longer wear would make them more comfortable | • Not comfortable<br>• Uncomfortable under 5<sup>th</sup> met<br>• Uncomfortable – can’t wear all day<br>• Discomfort at 5<sup>th</sup> MET/ lateral<br>• Feels like a met pad<br>• Minor irritation on left 1<sup>st</sup> met<br>• Discomfort on lateral arch region<br>• Pain in middle of foot<br>• Did not like arch height – arch felt sore<br>• Right foot 5<sup>th</sup> met hurts |
|  | *Supportiveness* |  |  |
|  |  | • Arch not supportive enough | • Missing support lateral left foot<br>• No arch support<br>• Arch too high |
|  | *Stability/ Balance* |  |  |
|  | • Reduced stability | • Less stable – but got used to it<br>• Less stable than no insole<br>• Reduced stability/ Felt wobbly | • More dramatic trunk lean<br>• Worse balance/ feels unstable<br>• Back on heels<br>• Unstable |
|  | *Materials* |  |  |
|  | • Not smooth – somewhat pliable | • Feels less bouncy, more firm, and more cupped in the heels<br>• Not soft enough/ not enough padding | • Feels firm/ Would like it to be softer<br>• Needs softening under arch<br>• Does not like the material |
|  | *Fit in the shoe* |  |  |
|  | • Medial heel feels elevated<br>• Right foot had to tilt to keep heel down | • Seems thicker – foot sitting higher in shoe | • Heel feels like it is slipping loose |
|  | *Misc.* |  |  |
|  | • Would not want – felt unnatural<br>• Clenching toes<br>• Pushes foot forward | • Toes pushed into front of shoe by arch height | • Pushing foot outward<br>• Supinating<br>• Weird gap leaning – foot limping<br>• Didn’t like too much<br>• Toes squish at front – uncomfortable<br>• Medial edge too high and pushing feet to lateral<br>• Causing tingling and burning |

#### Comfort/pain

Both the SoC and PB-CAI were reported as “comfortable”, while this word was never used to describe the FE-CAI. Both the SoC and FE-CAI were described as either “uncomfortable/not very comfortable” or “causing discomfort” in a specific location of the foot. The PB-CAI was not described in this way. Both the PB-CAI and FE-CAI were described as “not painful” and “painful [in a specific location of the foot]”.

#### Supportiveness

All three insoles were reported as “supportive”, particularly the PB-CAI and FE-CAI were “supportive in the arch”. However, the PB-CAI and FE-CAI were also reported as “not supportive enough” in specific regions of the foot. The SoC was not described as “supportive”.

#### Stability/balance

All three insoles were reported as both “stable” or “better balance” and “unstable” or “worse balance”. The SoC has more reports of “stable”, while the FE-CAI had more reports of “unstable”. The PB-CAI had similar reports of both “stable” and “unstable”.

#### Materials

The PB-CAI and FE-CAI were both reported to feel “firm” or to “not have enough padding”, while the SoC was indicated to have “cushion”.

#### Fit in the shoe

The SoC and PB-CAI were reported to “fit well in the shoe”. However, all three insoles were reported to have the “heel slipping out [of the shoe during walking]”.

## DISCUSSION

The aim of this study was to assess the effectiveness of two novel 3D-printed custom accommodative insoles, PB-CAI and FE-CAI, in reducing plantar pressure compared to the SoC and NI conditions in individuals with diabetes. The results provide strong evidence that the 3D-printed insoles offer superior plantar pressure reduction, particularly in regions of high pressure, compared to both the NI condition and the SoC insoles. Notably, while both PB-CAI and FE-CAI were effective at reducing plantar pressure, PB-CAI stood out in terms of comfort and participant preference, suggesting that the design and material properties of custom 3D-printed insoles could provide a more patient-centered solution for diabetes-related foot complications.

### Clinical Significance of Plantar Pressure Reduction

The reduction in plantar pressure, especially in regions at high risk of ulceration, is critical in preventing diabetic foot ulcers. The strategies employed by each insole design to reduce plantar pressure and their corresponding outcomes differ, warranting further discussion.

#### SoC

The SoC insoles, constructed using conventional methods and materials (e.g., EVA base with Plastazote-PORON top layer), demonstrated significant reductions in PPP and PTI in the ROI when compared to the NI condition (Figures 2A and 2C). This is contrary to our previous findings in healthy participants (11), but is consistent with previous studies evaluating the effects of the SoC on the intended population, individuals with diabetes with ulcer, neuropathy, or deformity (10,26–29). While SoC insoles were effective at reducing pressure in the ROI, they did not provide further reductions in the adjacent (ADJ) regions (Figures 2B and 2D), nor were they tailored to the patient-specific plantar pressure. Their effectiveness is likely attributed to the general offloading properties of the soft materials, but their uniformity limits precision pressure redistribution.

#### PB-CAI

The PB-CAI insoles were designed using in-shoe pressure data to identify localized high-pressure regions (≥200 kPa), and designed with a sparse lattice to accommodate these areas. This localized offloading approach effectively reduced both PPP and PTI in the ROI significantly beyond that of the SoC (Figures 2A and 2C). Due to this concentrated design feature, PB-CAI did not significantly increase PPP or PTI in the ADJ region. This is expected given that the design strategy focuses on offloading specifically in the ROI while maintaining structural integrity and support in surrounding areas. This may be beneficial in cases where pressure is confined to a small anatomical region, as it enables precise relief without broad redistribution, which could disrupt mobility or comfort. Consistent with previous findings in healthy adults, the PB-CAI did not significantly increase pressure in the ADJ region (11), lending further support to the targeted offloading on the ROI. Offloading in the ADJ may be less clinically meaningful as these regions are already below the ulceration threshold. Overall, PB-CAI provides targeted pressure relief, making it especially suitable for patients with discrete plantar pressure risks. Its success in reducing pressure without introducing new high-pressure zones in adjacent tissue suggests a safe and effective design for clinical use.

Several participants showed increases in PPP in the ADJ, although not significantly. We believe there is potential to improve the design of the offloading region, as outlined in future directions. Additionally, we acknowledge a limitation in the ability of current tools to accurately capture in-shoe plantar pressure in the same location on the foot and insole across multiple conditions, as described in the limitations section.

#### FE-CAI

In contrast to the PB-CAI, the FE-CAI employed a computational, simulation-based workflow using FE modeling to simulate forefoot loading and guide structural changes such as adjusting the metatarsal bar height. This method enabled a more global offloading strategy by considering broader forefoot mechanics during insole design.

The FE-CAI significantly reduced PPP and PTI in both the ROI and ADJ regions (Figure 2), the latter being a unique benefit among all insoles tested. This broader redistribution of load may explain why FE-CAI achieved the greatest overall PTI reduction. The FE-CAI’s use of a metatarsal bar appears to facilitate prolonged load distribution across the entire forefoot, rather than concentrating relief in a single area. This might be especially beneficial for patients with multiple high-risk zones or more diffuse forefoot loading, where a global pressure-relief strategy is warranted. While the additional offloading in the ADJ may be less clinically meaningful in regions already below ulceration thresholds, it still supports the insole’s effectiveness in minimizing tissue stress throughout the forefoot.

#### Summary

It is encouraging that none of the insoles significantly increased PPP or PTI in the ADJ region, suggesting that all insole conditions safely redistributed plantar pressure. The FE-CAI uniquely provided significant reductions in both ROI and ADJ, suggesting a broader protective effect, while PB-CAI delivered highly targeted pressure relief. The SoC, while effective, lacked the precision and adaptability of the 3D-printed options. The overall mean pressures were reduced in the 3D-printed insole conditions; however, there were several instances of in-shoe peak pressure of 200 kPa or more (19–21) in both the ROI and ADJ. We discuss the reasoning for this in the methods and limitations section. As described in the future work section, optimizing the transition between the offloading zone and surrounding areas, similar to (30), may help lower the elevated PPP observed in the ADJ. In summary, the pressure findings underscore the potential of personalized 3D-printed insoles to provide more precise pressure relief, which could play a significant role in reducing the incidence of diabetic foot ulcers.

### Participant Preferences

While the PB-CAI and FE-CAI demonstrated superior pressure relief, the subjective comfort and participant preferences are an additional takeaway from this study. Notably, 40% of participants preferred the PB-CAI, citing its comfort, support, and fit. Comfort, particularly in the arch region, was frequently mentioned as a positive attribute of PB-CAI, which might explain its higher preference compared to FE-CAI, even though the latter also effectively reduced plantar pressures. While FE-CAI was most effective in pressure relief, it was generally less favored by participants. The majority of the dislikes of the FE-CAI appear to be attributed to the comfort of the metatarsal bar. This is the key mechanism for offloading the ROI, but it appears to cause discomfort in some users.

All three insoles were reported to both improve and worsen stability/balance. While this subjective finding is inconsistent across study participants, it is not supported by previous quantitative findings of no systematic differences in standing balance across a subset (NI, SoC, and PB-CAI) of the insole conditions (31).

Overall, the subjective reports suggest that insole design, beyond its capacity to offload the areas of high pressure, is critical for the user experience. Factors such as fit, perceived support, and material should not be overlooked. Interestingly, despite being reported as less comfortable than the PB-CAI, the FE-CAI was still considered supportive, particularly in the arch area. There is a potential trade-off between the mechanical performance of the insole and its perceived comfort. Of note, the relationship between plantar pressure metrics and subjective perceptions such as comfort or supportiveness remains unclear. We speculate that a more uniform distribution of plantar pressure may not directly correlate with increased comfort. Different regions of the foot may be more accustomed to tolerating higher or lower pressures, and the pressure distribution and comfort may be user specific. Future work could focus on designing for perceived comfort in addition to offloading capacity or evaluating the relationships between pressure redistribution comfort and balance. This aligns with previous literature that emphasizes the importance of comfort and usability in ensuring patient compliance with insole interventions (32).

Research into 3D-printed custom accommodative insoles is rapidly expanding, suggesting that 3D-printed custom accommodative insoles are a promising approach to improve fit, support, and plantar pressure reduction in people with diabetes. These insoles can be individually tailored with patient-specific geometry (arch profile, metatarsal additions) and stiffness (pressure-informed design) to optimize offloading, especially in high-risk areas (33–37). Strategies such as graded-stiffness designs in the heel (30) and pressure-informed customization in the forefoot have shown superior performance over standard of care and traditional footwear (35). More recently, researchers have been developing prototypes of active insoles that employ soft robotics to modulate stiffness and minimize plantar pressure during walking (38). While these innovations are promising, most research, as does the current study, focuses on pressure reduction rather than direct ulcer prevention outcomes (39). The current study contributes to this growing body of work by incorporating metatarsal bars and selectively offloaded regions with softer lattice structures within an existing insole to enhance targeted pressure relief.

### Limitations

A limitation of this work is related to the tools used to evaluate plantar pressure. Specifically, a location shift in high-pressure sensels was sometimes observed on the second day of data collection. This is likely attributable to a logistical limitation of the study equipment. Specifically, the pedar insole is not secured to the foot or the shoe, allowing it to be in slightly different positions across conditions. Additionally, the foot can shift and move over the CAI, as a result, the pressure sensels and the ROI are not consistently aligned with the same regions of the foot or insoles across all conditions, leading to some participants showing peak pressures in slightly inconsistent locations. This explains some of the increased variability in ADJ for PB-CAI (and to a lesser extent FE-CAI and SoC), as a portion of participants have pressures > 200kPa in this region. Finally, we acknowledge that the pressure remains over the 200kPa threshold in the ADJ region in some instances of both 3D-printed insoles. We are working towards eliminating these unwanted secondary effects in future designs (for example, by exploiting the lattice construction to produce stiffness gradients around high-pressure regions, as mentioned in the future research section). We note that the SoC device also introduced these effects in the ADJ for a large number of subjects, suggesting that this is an inherent risk with the current approach to custom accommodative insole design. Our existing FE-CAI design significantly reduced these adjacent region effects compared to the standard of care, indicating we are already able to provide an improvement in pressure redistribution.

### Future Research

The findings of this study suggest several avenues for further exploration. First, while the 3D-printed insoles showed substantial biomechanical advantages over the SoC insoles, issues related to heel slippage in the shoe were reported across all insole types. This suggests that improvements in fit—especially in the heel area—for both the PB-CAI and FE-CAI could enhance the overall effectiveness of these custom insoles. Future designs will address this shoe fit issue to ensure functionality during dynamic activities like walking. Additionally, while our study focused on a short-term, controlled walking trial to evaluate PPP and PTI, the long-term effects of these 3D-printed insoles remain unknown. Future studies will investigate how prolonged use of PB-CAI and FE-CAI in the real world affects foot health outcomes, such as plantar pressure, the prevention of diabetic foot ulcers, changes in gait patterns, and overall mobility. Further, assessing how 3D-printed accommodative insoles influence plantar pressure during more dynamic activities—such as turning or stair climbing—will offer additional insight into their overall effectiveness. Another interesting avenue for future research is to assess how the structural properties of 3D-printed insoles—such as lattice density, material stiffness, and the overall design of offloading regions—affect clinical outcomes. While the PB-CAI effectively reduced plantar pressure within the ROI, its design could be improved to also alleviate pressure in the adjacent regions (ADJ). Specifically, optimizing the transition between the offloading zone and surrounding areas may help lower the elevated PPP observed in the ADJ, similar to (30). The current design features an abrupt change in stiffness at the ROI boundary, which could be reconfigured into a more gradual transition to promote better pressure redistribution.

## Conclusion

This study demonstrates that novel 3D-printed custom accommodative insoles, PB-CAI and FE-CAI, provide significant improvements in offloading high plantar pressures compared to the SoC and NI conditions in individuals with diabetes. Both PB-CAI and FE-CAI significantly reduced PPP and PTI in targeted regions of interest, with FE-CAI also showing unique benefits in adjacent areas. Importantly, participant preference and comfort ratings highlighted the PB-CAI as the preferred option, emphasizing the need for patient-centered design in clinical orthotic interventions. These findings support the clinical potential of personalized, 3D-printed accommodative insoles as a more precise and adaptable tool for the prevention of diabetic foot ulcers. However, further research is needed to address long-term wear effects, optimize offloading region transitions, and explore performance during more dynamic activities. By refining design features such as stiffness gradients, fit, and material feel, future iterations of 3D-printed insoles could enhance both biomechanical function and patient compliance, offering a promising step forward in diabetic foot care.

## Data Availability

All data produced in the present study may be available upon reasonable request to the authors

## Acknowledgements

We acknowledge Ellen Clarissimeaux and Leo Gagnon from the Department of Veterans Affairs for their contributions to data collection.

## Author Contributions and Guarantor Statement

Conceptualization BM, ST, WL

Data curation KN MV EL ST BM DH

Formal analysis KN DH EL ST BM

Funding acquisition BM, WL, ST

Investigation BM KN CC MV ANC DH EL

Methodology BM, ST, WL, KN, EL, MV

Project administration BM KN

Resources BM, WL, ST

Software ST, DH, KN, MV, EL

Supervision BM, WL, ST

Visualization: DH, KN, MV, BM

Writing – original draft BM

Writing – review & editing KN CC ANC DH EL WL ST

All authors approved the final version of the manuscript.

